# Interleukin-6 blockade for severe COVID-19

**DOI:** 10.1101/2020.04.20.20061861

**Authors:** Mathilde Roumier, Romain Paule, Matthieu Groh, Alexandre Vallée, Félix Ackermann, for the Foch COVID-19 Study Group

## Abstract

In the context of COVID-19 pandemic and growing tensions worldwide regarding healthcare facilities, there is an urgent need for effective treatments likely to reduce the crunch of ICU beds. Following the assumption by Mehta and colleagues who exhorted physicians to screen patients with severe COVID-19 for hyperinflammation and investigate immunomodulatory drugs in this setting, we relate our short-term - yet promising - experience regarding IL6 blockade with tocilizumab in 30 selected patients of less than 80 years of age, >5 days of prior disease duration, severe (i.e. requiring strictly over 6L/min of oxygen therapy) rapidly deteriorating (i.e. increase by more than 3L/min of oxygen flow within the previous 12 hours) COVID-19-related pneumonia. By comparison with a control group of patients (matched for age, gender and disease severity using the inverse probability of treatment weighted methodology) that did not receive tocilizumab. We demonstrate that, in highly selected patients, IL6 blockade could curb the “cytokine storm”, prevent ICU admission and the requirement for mechanical ventilation. Notwithstanding the shortcomings of this retrospective small sample-size study, we believe that these preliminary findings support the fostering of research efforts in the fight against COVID-19-induced inflammation, especially before patients require admission to the ICU.

---

The pathophysiology of Corona Virus Disease-19 (COVID-19) is still unclear. Yet, in a subgroup of patients with life-threatening acute respiratory distress syndrome (ARDS), there is growing evidence that virally-induced pro-inflammatory cytokines (including Interleukin (IL)1β, IL-6, tumor necrosis factor-α, and granulocyte colony stimulating factor) lead to both hyperinflammatory and procoagulatory states at a late-stage of the disease (1,2). Such findings are further corroborated by recent studies, which highlighted that high levels of C-reactive protein, IL-6 and D-Dimer upon admission were predictors of mortality (3,4). In their recent Comment, Mehta and colleagues exhorted physicians to screen patients with severe COVID-19 for hyperinflammation and suggested that immunosuppressive and/or immunomodulatory drugs should be investigated in this setting (5). Here, we relate our short-term - yet promising - experience regarding IL6 blockade for severe COVID-19 with tocilizumab.

Tocilizumab is mainly used worldwide for the treatment of rheumatoid arthritis, but other indications include refractory giant cell arthritis, idiopathic multicentric Castleman disease, and chimeric antigen receptor T-cell therapy-induced cytokine release syndrome (a condition that to some extent can mimic the clinical picture of COVID-19- induced ARDS) (6). In the emergency context of the COVID-19 epidemic in France, an off-label compassionate-use program of intravenous tocilizumab (8 mg/kg at the discretion of treating physicians, renewable once in case of insufficient response to therapy) was started at Foch university hospital (Suresnes, France) for selected COVID-19 patients (per WHO criteria) of less than 80 years of age with severe (*i*.*e*. requiring >6L/min of oxygen therapy) rapidly deteriorating (*i*.*e*. increase by ≥3L/min of oxygen flow within the previous 12 hours) pneumonia, high C-reactive protein levels and with ≥5 days of prior disease duration. All patients provided informed consent prior to receiving tocilizumab and this program was approved by the local Institutional Review Board (approval number: IRB00012437).

Between March 21^st^ 2020 and April 2^nd^ 2020, 30 patients (80% males; median age: 50 years) were treated with tocilizumab, including 7 (23%) in the setting of Intensive Care Unit (ICU) **(Table 1)**. After a median (IQR) follow-up of 8 (6.0-9.75) days, and by comparison with a control group of patients (matched for age, gender and disease severity using the inverse probability of treatment weighted methodology with SAS software) (7) that did not receive tocilizumab, IL-6 blockade significantly reduced the requirement of subsequent mechanical ventilation (weighted OR: 0.42; 95%CI[0,20-0,89]; p=0,025). Although unadjusted analysis showed a trend towards a reduction of mortality (OR: 0.25 95%CI [0.05-0.95], p=0.04), statistical significance disappeared after weighted analysis. Last, when considering only 23 patients (and 16 controls) treated outside the ICU, treatment with tocilizumab significantly reduced the risk of subsequent ICU admission (weighted OR: 0.17; 95%CI[0,06-0,48]; p=0,001). Overall, as of April 4^th^ 2020, of the 30 patients treated with tocilizumab, 3 (10%) had died, while 4/7 (57%) and 6/30 (20%) were discharged from the ICU and from hospital, respectively. Overall, tocilizumab was well-tolerated, yet mild hepatic cytolysis (n=2) and ventilator-acquired pneumonia (n=1) were reported. Of note, 2 patients treated with tocilizumab also received a 10-day course of hydroxychloroquine (200mg tid) and azithromycin (250mg bid on day1 and qid thereafter), while 2 patients (that were not treated with tocilizumab) received high-dose (≥1mg/kg/d) methylprednisolone pulses.

**Table 1:** Demographic, clinical, laboratory, radiographic findings and outcomes of the overall population.

|  | Unadjusted population |  |  | Weighted Population |  |  |
| --- | --- | --- | --- | --- | --- | --- |
|  | Treated<br>N=30 | Untreated<br>N=29 | P-value | Treated<br>N=30 | Untreated<br>N=29 | P-value |
| **Outcomes** |  |  |  |  |  |  |
| Invasive ventilation | 10 (33.3) | 16 (55.2) | 0.089 | 43.1% | 64.0% | 0.025 |
| Death | 3 (10.0) | 9 (31.0) | 0.041 | 17.2% | 18.7% | 0.837 |
| **Parameters used for the propensity score** |  |  |  |  |  |  |
| Age | 58.8 (12.4) | 71.2 (15.4) | 0.001 | 62.3 (11.3) | 60.6 (21.6) | 0.598 |
| Female | 6 (20.0) | 6 (20.7) | 0.947 | 21.9% | 16.4% | 0.459 |
| Baseline ICU hospitalization | 7 (23.3) | 13 (44.8) | 0.079 | 33.4% | 50.3% | 0.066 |
| COPD | 4 (13.3) | 7 (24.1) | 0.284 | 15.1% | 16.0% | 0.888 |
| Diabetes | 7 (23.3) | 10 (34.5) | 0.343 | 29.5% | 27.6% | 0.819 |
| Obesity | 5 (16.7) | 4 (13.8) | 0.759 | 19.6% | 31.9% |  |
| NEWS 2 score |  |  | 0.786 |  |  | 0.734 |
| score ≥ 7 | 24 (80.0) | 24 (82.8) | | 83.3% | 85.6% | |
| score 5-6 | 6 (20.0) | 5 (17.2) |  | 16.7% | 14.4% |  |
| C-reactive protein (mg/L) | 189.0<br>(104.4) | 167.4<br>(106.8) | 0.426 | 186.7<br>(112.7) | 160.8<br>(93.7) | 0.179 |
| Eosinopenia | 21 (70.0) | 25 (86.2) | 0.129 | 78.8% | 88.7% | 0.146 |
| Time between hospitalization and the need for > 6L/min of oxygen therapy (hours) | 1.8 (3.1) | 4.1 (11.8) | 0.263 | 1.5 (2.6) | 2.4 (8.5) | 0.428 |
| **Comorbidities** |  |  |  |  |  |  |
| Hypertension | 6 (20.0) | 18 (62.1) | 0.008 |  |  |  |
| Cardiovascular disease | 4 (13.3) | 10 (34.5) | 0.053 |  |  |  |
| Cerebrovascular disease | 0 (0.0) | 3 (10.3) | 0.071 |  |  |  |
| Chronic kidney disease | 4 (13.3) | 5 (17.2) | 0.676 |  |  |  |
| HIV | 2 (6.7) | 1 (5.6) | 0.877 |  |  |  |
| Immunosuppressive therapy | 4 (13.3) | 3 (15.0) | 0.868 |  |  |  |
| **Clinical findings** |  |  |  |  |  |  |
| BMI (kg/m<sup>2</sup>) | 27.6 (3.8) | 25.5 (3.9) | 0.183 |  |  |  |
| Time of first symptoms to Tocilizumab onset (days) | 14.1 (3.5) | .. | .. |  |  |  |
| **Paraclinical findings** |  |  |  |  |  |  |
| Positive PCR for SARS-CoV2 | 29 (96.7) | 29 (100) | 1.000 |  |  |  |
| Lymphopenia | 23 (82.1) | 17 (85.0) | 0.793 |  |  |  |
| Ferritin (μg/L) | 2682.8<br>(3753.2) | 1827.9<br>(1221.2) | 0.391 | | | |
| D-dimer (mg/L) | 3712.8<br>(5836.4) | 1880.5<br>(2162.7) | 0.335 |  |  |  |
| ≥ 50% lung involvement on CT-scan | 18 (85.7) | 6 (85.7) | 1.000 | | | |

Data are n (%) and mean (SD). P-values were calculated using Mann-Whitney U test, chi-square test or Fisher's exact test, as appropriate. COPD = chronic obstructive pulmonary disease. CoV = Coronavirus. CT = computed tomography. ICU= Intensive Care Unit. NEWS= National Early Warning Score. PCR = polymerase chain reaction. SARS = Severe Acute Respiratory Syndrome.

In the context of COVID-19 pandemic and growing tensions worldwide regarding healthcare facilities, there is an urgent need for effective treatments likely to reduce the crunch of ICU beds. Concordant with the assumption by Mehta (5) and others (8), these data suggest that targeting IL-6 in highly selected patients with rapidly deteriorating pneumonia and high inflammatory parameters could curb the “cytokine storm”, prevent ICU admission and the requirement for mechanical ventilation. These results are in line with the encouraging data reported by Xu and colleagues regarding the use of tocilizumab in COVID-19 (9), but differ from those of Gritti and colleagues who treated more severe patients requiring non-invasive ventilation with siltuximab (another IL-6R-targeted therapy) (10). Notwithstanding the shortcomings of these retrospective small sample-size studies, we believe that these preliminary findings support the fostering of research efforts in the fight against COVID-19-induced inflammation, especially before patients require admission to the ICU. Evidently, the results of ongoing larger-scale prospective randomized-controlled trials evaluating tocilizumab (ChiCTR2000029765; NCT04320615), sarilumab (NCT04315298; NCT04324073) and siltuximab (NCTNCT04329650) are eagerly awaited. Moreover, other immunomodulatory treatment strategies including high-dose corticosteroids, IL-1 blockade or JAK inhibition also deserve to be investigated.

## Data Availability

all original data are available

